# ChatGPT Influence on Medical Decision-Making, Bias, and Equity: A Randomized Study of Clinicians Evaluating Clinical Vignettes

**DOI:** 10.1101/2023.11.24.23298844

**Authors:** Ethan Goh, Bryan Bunning, Elaine Khoong, Robert Gallo, Arnold Milstein, Damon Centola, Jonathan H Chen

**Author notes:** Corresponding author (Ethan Goh).

## Abstract

In a randomized, pre-post intervention study, we evaluated the influence of a large language model (LLM) generative AI system on accuracy of physician decision-making and bias in healthcare. 50 US-licensed physicians reviewed a video clinical vignette, featuring actors representing different demographics (a White male or a Black female) with chest pain. Participants were asked to answer clinical questions around triage, risk, and treatment based on these vignettes, then asked to reconsider after receiving advice generated by ChatGPT+ (GPT4). The primary outcome was the accuracy of clinical decisions based on pre-established evidence-based guidelines. Results showed that physicians are willing to change their initial clinical impressions given AI assistance, and that this led to a significant improvement in clinical decision-making accuracy in a chest pain evaluation scenario without introducing or exacerbating existing race or gender biases. A survey of physician participants indicates that the majority expect LLM tools to play a significant role in clinical decision making.

---

The emergence of large language model (LLM) (e.g., GPT-4 and Med-PaLM2) generative AI systems challenge the very nature of medical practice and education^1^ when these automated systems demonstrate surprising accuracy on medical exam questions vs. human benchmarks^2^. Yet these systems still remain unfit for autonomous medical decision making, given their propensity for confabulation, inconsistent behavior, lack of regulatory oversight, and the risk of unintended consequences such as exacerbating biases against underrepresented minority patients^3,4,5^. Rather than asking how an automated system alone performs on a medical exam, here we evaluate the more important and relevant question of whether a human practitioner can be augmented with AI support to improve clinical decision-making, and whether doing so will introduce or exacerbate undesirable bias observed in prior studies on individual vs. collective clinical decision making^6^.

Participants reviewed a video clinical vignette of a standardized patient complaining of chest pain (Figure 1), with participants randomly assigned to have the case feature a White male or a Black female as used in a previous study^6^ demonstrating human biases in clinical interpretation. Participants then answered four multiple choice clinical questions based on these vignettes (full case material and questions in Supplementary Materials 2 and 3), with the option of using any information resource available (e.g., MDCalc, Up-to-Date, PubMed) *except* for LLM AI systems. Participants were then allowed to review answer suggestions generated by ChatGPT+ (GPT-4 used between May to August 2023), and subsequently given the option to modify their original answers.

Table 1 reports the participants’ average scores on the clinical questions in each arm of randomization (White male patient vs. Black female patient) before and after exposure to GPT-4 responses. A statistical model showed significant differences in scores between the groups and pre-vs. post-LLM (Supplementary 6 and 7).

**Table 1:** Clinician Decision Accuracy Before and After LLM Intervention.

|  |  |  |  |
| --- | --- | --- | --- |
| **Average Score Metric** | **Group A (White Male) N=25** | **Group B (Black Female) N=25** | **Absolute Difference between A and B** |
| Initial (Out of 4) | 1.9 (47%) | 2.5 (63%) | 0.62 (16%) |
| Post-LLM (Out of 4) | 2.6 (65%) | 3.2 (80%) | 0.60 (15%) |
| Pre-Post Change (Out of 4) | +0.72 (18%) | +0.70 (18%) | -0.02 (-1%) |

These results indicate that physicians are willing to change their clinical decisions based on responses from an automated large language model AI system, as opposed to anchoring on their initial decisions and skeptically refusing to be swayed by a computer-based response^7^. Moreover, doing so significantly improved the accuracy of their answers in this scenario.

A previous study^8^ established the validity of the clinical vignette and standardized patient videos, while also demonstrating bias in physician answers that could be mitigated through a crowdsourcing process. In contrast to that previous study, our statistical model, which adjusted for group and pre/post score, found that participants were more accurate when viewing the Black female video vs. the White male video (p < 0.01) (Supplementary 6). The reason for the differing result is unclear, but could perhaps be attributed to the Hawthorne effect^9^, as participants completed this study in a virtual meeting setting while being observed by a researcher. In either case, our statistical model (Supplementary 6) showed a significant improvement in participant scores post-intervention (p < 0.000001). This improvement was achieved without introducing or exacerbating any race or gender biases.

Different question types (triage, risk assessment, and treatment) were based on the previously established study and selected to mirror the variation in real-world clinical decisions that physicians encounter. Having a range of question types that involve judgment skills (risk and triage) vs. knowledge-base (evidence-based treatment selection) allowed us to assess the potential differential impact of potential bias and AI interaction methods on physician decision-making. Having a prepared LLM-response for support in questions #1 and #2 ensured consistency in the user interaction, while the participant’s free open-ended use of ChatGPT+ for question #3 and #4 allowed for additional qualitative analysis of the types of queries and interactions physicians would have with such a system in a live setting. Breakdown of the question accuracy results are summarized in Supplementary 5.

Table 2 describes categories of participant interactions with the AI chatbot when they were allowed freeform interaction with ChatGPT+ for treatment selection in questions #3 and #4, illustrating the multifaceted relevance of such technology in clinical decision-making settings. The usage patterns range from seeking clarification on guidelines and evidence-based practice to soliciting advice on specific patient scenarios. Specific examples of the participant’s chat log with the LLM are included, illustrating that many directly copy-pasted the clinical vignette or question content into the chat interface, while others asked further probing or summarized questions. While these findings are context-specific, they provide an initial understanding of the different types of physician / AI chatbot interactions and potential applications in clinical decision processes.

**Table 2:** Categorization of Participant Interactions with ChatGPT in a Chest Pain Clinical Vignette.

|  |  |  |
| --- | --- | --- |
| Interaction | Description | Chat Log Transcript Excerpt |
| 1. Guideline and Evidence-based Practice Discussions | Physicians engaged with the AI to inquire about evidence supporting clinical decisions. This encompassed both specific clinical trials and current guidelines. | "What are the American Heart Association recommendations for a patient presenting with a NSTEMI and a TIMI score of 4 for initial medical therapy?"<br>"What medical interventions have been shown to improve outcomes for NSTEMI?" |
| 2. Patient Scenario-Based Inquiries | Physicians presented patient details to the AI, asking for advice or confirmation on management strategies. | “For a patient with chest pain, ST depressions, troponin elevations, what is optimal management?”<br>"I have a 65 year old patient with a history of hypertension and recent dyspnea on exertion who is now experiencing worsening chest pain, elevated troponin and ST depressions on their EKG. There were no signs of heart failure or cardiogenic shock. In addition to potential catheterization, which of the following medical interventions are most likely to improve the patient's clinical outcome?" |
| 3. Validation of Own Knowledge or Beliefs | Physicians used the AI to validate or challenge their own clinical knowledge or beliefs, especially by asking follow-up questions to clarify or expand on the information provided. | “I believe that nifedipine is contraindicated due to reflex sympathetic activation, is this correct?”<br>“Why is sacubitril/valsartan not the right answer?”<br>“Why did you not include ARNIs?” |
| 4. Exploring<br>New or<br>Emerging<br>Treatment<br>Modalities | Physicians inquired about<br>new or less familiar<br>treatments and their benefits<br>in specific clinical scenarios. | "What is the evidence supporting empagliflozin for<br>post-MI?"<br>"Does empagliflozin reduce mortality in patients<br>with CAD after PCI?" |
| 5. Comparative<br>Analysis of<br>Treatment<br>Options | Physicians used the AI to<br>analyze and compare<br>different treatment options. | "Can you list your reasoning for the correct answer:<br>A. [list of medications] B. [list of medications]...?"<br>"Are ACE inhibitors used in an acute NSTEMI?"<br>"Are calcium channel blockers recommended in the<br>acute management of patients with acute coronary<br>syndrome?" |

90% of participants indicated in a post-task survey (Supplementary 8 and 9) that large language model tools like ChatGPT will play a significant role in healthcare for clinical decision making, with 66% rating the likelihood as ‘very likely’ and 24% as ‘somewhat likely’. Recommendations for improving the utility of AI chatbots in healthcare were varied but focused on increasing clinical relevance, such as by developing a healthcare-specific user interface and enhancing its ability to process and interpret patient information. Transparency in AI reasoning and decision-making processes was also a significant concern, with a call for AI chatbots to provide evidence-based citations for its recommendations.

A limitation of this study design is that the physician participants were given video of a standardized patient case and an ECG image to review, whereas ChatGPT+ at the time of the study only allowed for text interaction, requiring it to be given a text-based summary of the clinical vignette. Moreover, such LLMs exhibit variation in their outputs based on prompt variations, algorithmic updates, and underlying randomness. We respectively developed the vignette content with varying case prompts (including with vs. without demographic information) to confirm that the produced LLM outputs remained similar in meaning (and not showing different suggestions depending on the patient’s stated race or gender that has been observed under other specific adversarial scenarios)^10^.

The present study was limited to a single clinical vignette to isolate the human-computer interaction phenomenon, and is not intended to represent the broad scope of medical practice. This case vignette was established for rigorous evaluation in a previous study^6^ to assess for biases through video-recorded standardized patients and evidence-based reference answer evaluations. While further evaluations can be done across a broader set of cases, this study represents a critical milestone that moves beyond the many studies evaluating LLM (vs. human) performance on medical questions to directly administering, observing, and evaluating the interaction and impact of augmenting human physicians with cutting edge LLM generative AI systems.

The results of this study indicate that large language model AI systems can significantly augment medical decision-making in a cardiac vignette, improving accuracy without introducing or exacerbating existing demographic bias. Interactions between clinicians and the AI chatbot indicate that physicians are receptive to the AI-based suggestions with potential usefulness across a spectrum of clinical question types.

## METHODS

We employed a randomized pre-post intervention design, approved by Stanford University’s IRB, to assess the impact of AI-assisted decision-making in healthcare. 50 US-licensed physicians with experience in Family Medicine, Internal Medicine, or Emergency Medicine were enlisted (Supplementary 1). Participants reviewed a video of an actor portraying a clinical scenario of chest pain and associated ECG results developed for a previous study on clinical bias^6^ (Supplementary 2). Participants were randomized to observe a White male or a Black female actor, and then tasked with responding to multiple choice clinical questions on immediate care triage, risk assessment, and medication management (Supplementary 3). Participants were permitted to use any external resources typically employed in their daily work, such as MDCalc, PubMed, and UpToDate. Subsequently, participants reviewed ChatGPT+ (GPT-4) generated responses from April 2023 (Supplementary 4) based on the case vignette information for questions #1 and #2, or directly interacted with ChatGPT+ for assistance for questions #3 and #4. Participants were given the option to change their answers after the above information intervention. The primary outcome measure was accuracy of answers to the clinical decision questions, based on evidence-based literature review^6^. As a secondary measure, we studied the variance in accuracy before and after intervention between both groups.

We analyzed results using R (v4.1.2) with a pre-specified Linear Mixed Effects Model (LMM) using the LME4 package (v1.1-34), with a random intercept for each participant. The model was first structured as: “Score (#correct out of 4 questions) ∼ pre/post-recommendation + experimental-group + interaction-term + (1|participant)” with binary covariates. After modeling, the interaction term did not significantly improve the model (ANOVA, p=0.88), and was dropped. Reported characteristics are from the LMM without an interaction term. The reference of the model covariates are pre-intervention and Group A (White male). Scores were treated as continuous variables. Model values were assessed at an unadjusted significance threshold of alpha=0.05 using Satterthwaite’s t-test. Pre-study power calculations were done to estimate adequate sample size and plan for adequate recruitment.

Following the completion of the clinical tasks, participants were asked to complete a survey to assess their perceptions of LLM tools like ChatGPT in healthcare (Supplementary 8). All participant interactions with ChatGPT+ (i.e., chat logs) were recorded and coded using an inductive qualitative data analysis approach to identify emergent themes^11,12^. This process was iterative, allowing categories to be refined for a precise representation of the interactions. E.G. independently coded the transcripts through readings of the transcripts. R.G. reviewed all transcripts subsequently to validate the coding.

**Figure 1:**
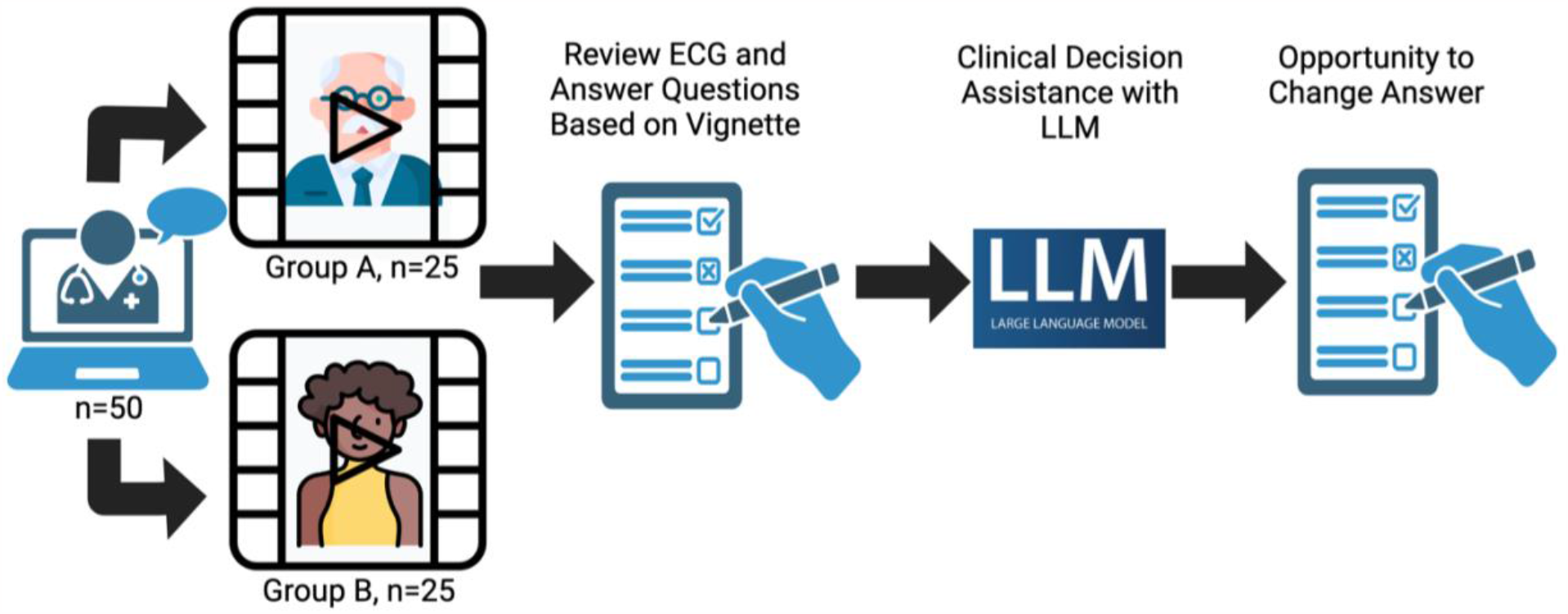
- Study Design: 50 US-licensed physicians were recruited for a remote video session where they were presented with a video of a standard patient actor depicting a case of chest pain in an outpatient setting. Participants were randomized to encounter an actor who was a White male or a Black female. The clinicians then responded to a series of four questions based on the vignette. For the first two questions, after providing their initial answers, they were presented with a pre-prepared LLM response based on the same vignette and questions. Clinicians were then offered an opportunity to modify their initial answers. For the final two questions, after their initial response, clinicians were allowed to directly interact with the LLM to ask any questions before considering whether or not to modify their answer.

## Data Availability

All LLMs outputs are included in the supplement with the prompts used. Transcript chat logs, raw score table, and individual survey responses are available upon request.

## Data Availability

All data produced in the present study are available upon reasonable request to the authors.

## CONTRIBUTIONS

Ethan Goh - Study design, data acquisition, data interpretation, manuscript preparation

Bryan Bunning - Study design, data interpretation, statistical analysis, critical revision

Elaine Khoong - Study design, technical and material support, data interpretation, critical revision

Robert Gallo - Data analysis, data interpretation, critical revision

Arnold Milstein - Study design, data analysis, data interpretation, funding and administrative support, critical revision

Damon Centola - Study design, technical and material support

Jonathan Chen - Study design, data analysis, data interpretation, critical revision, supervision, funding and administrative support

## Acknowledgements

Study design - Manoj Maddali, Matthew Schwede, Honor Magon, Isabelle Tan, Angel Arnault, Marissa Chua

## Affiliations, Disclosures and Funding

### Ethan Goh, MD, MS

Affiliations

- Stanford Center for Biomedical Informatics Research, Stanford University, Stanford, California, USA
- Stanford Clinical Excellence Research Center, Stanford University, Stanford, California, USA

Disclosures

- None

Funding

- None

### Bryan Bunning, MS

#### Affiliations

- Stanford Department of Biomedical Data Science. Stanford University, Stanford, California USA
- Quantitative Sciences Unit, Division of Biomedical Informatics Research, Department of Medicine, Stanford University School of Medicine, Stanford, CA, USA

#### Disclosures

- None Funding
- National Library of Medicine (2T15LM007033)

### Elaine Khoong, MD, MS

#### Affiliations

- Division of General Internal Medicine at San Francisco General Hospital, Department of Medicine, University of California San Francisco
- UCSF Center for Vulnerable Populations at San Francisco General Hospital Disclosures
- None Funding
- National Heart Lung and Blood Institute of the NIH under Award Number K23HL157750.

### Robert Gallo, MD

#### Affiliations

- Center for Innovation to Implementation, VA Palo Alto Health Care System Disclosures
- None Funding
- Dr. Gallo is supported by a VA Advanced Fellowship in Medical Informatics. The views expressed are those of the authors and not necessarily those of the Department of Veterans Affairs or those of the United States government.

### Arnold Milstein, MD

#### Affiliations

- Stanford Clinical Excellence Research Center, Stanford University, Stanford, California, USA Disclosures
- Dr Milstein reported uncompensated and compensated relationships with care.coach, Emsana Health, Embold Health, EZPT, FN Advisors, Intermountain Healthcare, JRSL, The Leapfrog Group, Peterson Center on Healthcare, Prealize Health, PBGH

#### Funding

- Pooled philanthropic gifts to Stanford University
- Research funding from Stanford Healthcare and Stanford Childrens Health

### Damon Centola, PhD

#### Affiliations

- Communication, Sociology and Engineering, University of Pennsylvania, PA Disclosures
- None

#### Funding

- DC gratefully acknowledges support from a Robert Wood Johnson Pioneer Grant.

### Jonathan H. Chen, MD, PhD

#### Affiliations

- Stanford Center for Biomedical Informatics Research, Stanford University, Stanford, California, USA
- Division of Hospital Medicine, Stanford University, Stanford, California, USA
- Stanford Clinical Excellence Research Center, Stanford University, Stanford, California, USA

#### Disclosure

- Co-founder of Reaction Explorer LLC that develops and licenses organic chemistry education?software.
- Paid consulting fees from Sutton Pierce, Younker Hyde MacFarlane, and Sykes McAllister as a medical expert witness.

#### Funding

- NIH/National Institute of Allergy and Infectious Diseases (1R01AI17812101)
- NIH/National Institute on Drug Abuse Clinical Trials Network (UG1DA015815 - CTN-0136)
- Gordon and Betty Moore Foundation (Grant #12409)
- Stanford Artificial Intelligence in Medicine and Imaging - Human-Centered Artificial Intelligence (AIMI-HAI) Partnership Grant
- Doris Duke Charitable Foundation - Covid-19 Fund to Retain Clinical Scientists (20211260)
- Google, Inc. Research collaboration Co-I to leverage EHR data to predict a range of clinical outcomes.
- American Heart Association - Strategically Focused Research Network - Diversity in Clinical Trials

