## Supplementary Materials for "ChatGPT Influence on Medical Decision-Making, Bias, and Equity: A Randomized Study of Clinicians Evaluating Clinical Vignettes"

| Supplementary 1: Demographics | | | |
|---|---|---|---|
| **Characteristic** | **Group A (White Male Vignette) N=25** | **Group B (Black Female Vignette) N=25** | **Total (N=50)** |
| **Gender** | | | |
| Male | 15 (60%) | 17 (68%) | 32 (64%) |
| Female | 10 (40%) | 8 (32%) | 18 (36%) |
| **Specialty** | | | |
| Family Medicine | 4 (16%) | 4 (16%) | 8 (16%) |
| Internal Medicine | 19 (76%) | 14 (56%) | 33 (66%) |
| Emergency Medicine | 2 (8%) | 7 (28%) | 9 (18%) |
| **Years of experience including residency training** | | | |
| Less than 3 years | 18 (72%) | 14 (56%) | 32 (64%) |
| 3 to 5 years | 3 (12%) | 3 (12%) | 6 (12%) |
| 5 to 10 years | 4 (16%) | 5 (20%) | 9 (18%) |
| More than 10 years | 0 (0%) | 3 (12%) | 3 (6%) |
| **Primary place of work** | | | |
| Academic medical center | 19 (76%) | 17 (68%) | 36 (72%) |
| Community hospital | 5 (20%) | 4 (16%) | 9 (18%) |
| Private practice (solo or group) | 1 (4%) | 3 (12%) | 4 (8%) |
| Multi-specialty group | 0 (0%) | 1 (4%) | 1 (2%) |

**Supplementary 2: Patient Script and ECG**

*Patient actor script:*

I'm so glad you were able to see me this afternoon. Ever since I retired a few years ago at 65, I've had time to try to get healthier. I know I'm overweight, so I've started to exercise more. After my walk this morning, I noticed a weird, tired feeling that made me feel a little short of breath. I sat down in my kitchen to get a sip of water and rest; it felt better a few minutes afterwards. I also felt fine when I walked up the stairs to your office. The medical assistant who took my vital signs said everything looks great, and I've been taking the blood pressure and cholesterol medication every day. So, I don't think it's a big deal, but I want to make sure since my dad had a heart attack in his early 60's.

*ECG exhibit:*

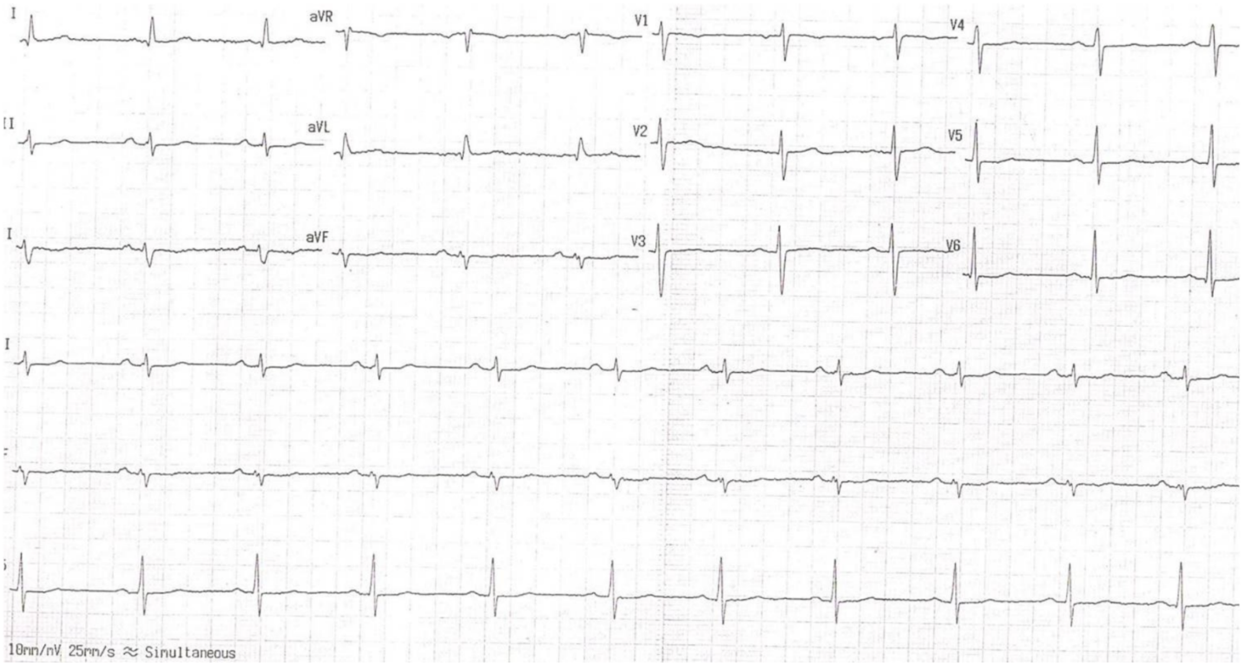

**Supplementary 3: Quiz questions and answers**

**Question 1: What would be your next step in management of this patient?**

a. Start a daily baby aspirin and provide clear return precautions. Schedule the patient to return in one week.
b. Start a daily baby aspirin and refer the patient for an urgent stress test within 2-3 days.
c. Provide a full-dose aspirin and refer to the emergency department for evaluation and monitoring.
d. Provide a full-dose aspirin and contact cardiology for urgent cardiac catheterization.

**Correct Answer: C** full-dose aspirin and refer to the emergency department.

Based on several risk stratification scores, this patient is at intermediate/moderate risk due to these components:

- symptoms (discomfort with exertion, dyspnea)
- history (concern for cardiac origin)
- age (>65 years old)
- EKG (T-wave inversion / flattening)
- risk factors (hypertension, hyperlipidemia, obesity, positive family history)

Specifically, the patient has a HEART score of 6 (1 point for moderately suspicious history; 1 point for repolarization disturbance; 2 points for age >65; 2 points for 3+ risk factors ) without a troponin level. Even a mild troponin increase would place the patient at 7 points (or high risk). Intermediate risk individuals were found to have a 12 – 16.6 % risk of a major adverse cardiac event (MACE) within 30 days and high-risk individuals had a 50-65% risk of a MACE. Without knowledge of a troponin level, this moderate-risk individual was further evaluated and admitted to the hospital in the HEART study.

A study within primary care created a risk score to determine the likelihood of coronary artery disease in patients with chest pain. From that study, the patient has a risk score of 3 (age 65+; worse during exercise; concern about cardiac origin) and potentially 4 (if pain is not reproducible by palpation). Patients with a risk score of 3+ were found to have 4.5 times likelihood of having coronary artery disease. The recommendation for this patient who also has T-wave abnormalities is for same day troponin testing or further evaluation in the emergency department.

Given both these studies, the patient needs to be immediately evaluated for further risk stratification via cardiac enzymes or a same day noninvasive stress testing, and therefore option C is the correct answer. Option A does not pursue necessary further evaluation. Option B delays this evaluation. Option D is not appropriate for an individual with intermediate risk.

*Reference:*


Bosner S, Haasenritter J, Becker A, Karatolios K, Vaucher P, Gencer B, et al. Ruling out coronary artery disease in primary care: development and validation of a simple prediction rule. Can Med Assoc J. 2010;182:1295–300.

Ebell MH. Evaluation of chest pain in primary care patients. Am Fam Physician. 2011;83:603–5.

Poldervaart JM, Reitsma JB, Backus BE, Koffijberg H, Veldkamp RF, ten Haaf ME, et al. Effect of Using the HEART Score in Patients With Chest Pain in the Emergency Department: A Stepped-Wedge, Cluster Randomized Trial. Ann Intern Med. 2017;166:689–697.


**Question 2: With the current information (and using risk classification tools such as the HEART score), what is this individual's chance of having a major adverse cardiac event within the next 30 days?**

a. <1%
b. 2 - 6%
c. 12 - 17%
d. 50 - 65%

**Correct Answer: C** 12 – 17 % within 30 days based on a HEART score of 6 (1 point for moderately suspicious history; 1 point for repolarization disturbance; 2 points for age >65; 2 points for 3+ risk factors ) without a troponin level.

*Reference:*

Mahler, S. A., Riley, R. F., Hiestand, B. C., Russell, G. B., Hoekstra, J. W., Lefebvre, C. W., ... & Herrington, D. M. (2015). The HEART pathway randomized trial. Circulation: Cardiovascular Quality and Outcomes, 8(2), 195-203.

**Question 3:** The same patient had worsening chest pain later in the day, and on evaluation was found to have elevated troponin and ST depressions on their EKG. There were no signs of heart failure or cardiogenic shock. In addition to potential catheterization, which of the following medical interventions are most likely to improve the patient's clinical outcome?

a. Aspirin, Heparin, Nitroglycerin, Supplemental oxygen
b. Aspirin, Calcium antagonists (e.g., nifedipine), Morphine, Nitroglycerin, Supplemental oxygen, Statin (e.g., atorvastatin)
c. ACE inhibitors (e.g., lisinopril), Aspirin, Calcium antagonists (e.g., nifedipine), Morphine, Nitroglycerin
d. ACE inhibitors (e.g., lisinopril), ADP receptor inhibitors (e.g., clopidogrel), Aspirin, Beta-blockers (e.g., metoprolol), Statin (e.g., atorvastatin)

**Correct Answer: D**

*Reference:*
Virani S, Newby L, et al. 2023 AHA/ACC/ACCP/ASPC/NLA/PCNA Guideline for the Management of Patients With Chronic Coronary Disease. J Am Coll Cardiol. 2023 Aug, 82 (9) 833–955.

**Question 4:** After inpatient treatment and stabilization, the patient has been discharged and is being reviewed in a cardiology outpatient clinic several months later.

Which of these medications have been shown to reduce mortality for patients with stable chronic ischemic heart disease?

a. ADP receptor inhibitors (e.g., clopidogrel), Aspirin, Beta-blockers (e.g., metoprolol, carvedilol), Statins (e.g., atorvastatin, rosuvastatin), Sacubitril/Valsartan (neprilysin inhibitor)
b. Anti-Arrhythmics (e.g., amiodarone), Anti-coagulants (e.g., warfarin, rivaroxaban, apixaban), Calcium Channel Blockers (e.g., amlodipine), Nitrates (e.g., isosorbide dinitrate), Oxygen supplementation

c. Beta-blockers (e.g., metoprolol, carvedilol), Loop Diuretics (e.g., furosemide), Implantable Cardioverter Defibrillator (ICD), Mineralocorticoid Receptor Antagonists (e.g., spironolactone)
d. ADP receptor inhibitors (e.g., clopidogrel), Angiotensin-Converting Enzyme (ACE) inhibitors / Angiotensin Receptor Blockers (ARBs) (e.g., lisinopril, losartan), Aspirin, Beta-blockers (e.g., metoprolol, carvedilol), Empagliflozin, Statins (e.g., atorvastatin, rosuvastatin)

**Correct Answer: D**

*Reference:*

Virani S, Newby L, et al. 2023 AHA/ACC/ACCP/ASPC/NLA/PCNA Guideline for the Management of Patients With Chronic Coronary Disease. J Am Coll Cardiol. 2023 Aug, 82 (9) 833–955.

**Supplementary 4: GPT-4 Prompt and Responses for Question 1 and Question 2**

| | **GPT-4 prompt** | **GPT-4 response** |
|---|---|---|
| **Question 1** | "I'm so glad you were able to see me this afternoon. Ever since I retired a few years ago at 65, I've had time to try to get healthier. I know I'm overweight, so I've started to exercise more. After my walk this morning, I noticed a weird, tired feeling that made me feel a little short of breath. I sat down in my kitchen to get a sip of water and rest; it felt better a few minutes afterwards. I also felt fine when I walked up the stairs to your office. The medical assistant who took my vital signs said everything looks great, and I've been taking the blood pressure and cholesterol medication every day. So, I don't think it's a big deal, but I want to make sure since my dad had a heart attack in his early 60's."<br><br>ECG showed T wave inversion/flattening.<br><br>Question 1: What would be your next step in management of this patient?<br><br>a. Start a daily baby aspirin and provide clear return precautions. Schedule the patient to return in one week.<br>b. Start a daily baby aspirin and refer the patient for an urgent stress test within 2-3 days. | If the ECG showed T wave inversion or flattening, this could potentially indicate underlying cardiac issues such as ischemia, which would warrant further investigation.<br><br>In this case, the appropriate next step would be: C. Provide a full-dose aspirin and refer to the emergency department for evaluation and monitoring.<br><br>This approach allows for a more thorough assessment of the patient's condition in a timely manner. The emergency department can perform further diagnostic testing, such as cardiac enzymes and additional imaging studies, to better understand the cause of the patient's symptoms and to rule out an acute coronary syndrome or other serious cardiac conditions. Additionally, the patient can be monitored in the emergency department to ensure their safety in case their condition worsens. |

| | | |
|---|---|---|
| | c. Provide a full-dose aspirin and refer to the emergency department for evaluation and monitoring.<br>d. Provide a full-dose aspirin and contact cardiology for urgent cardiac catheterization. | |
| **Question 2** | Question 2: With the current information (and using risk classification tools such as the HEART score), what is this individual's chance of having a major adverse cardiac event within the next 30 days?<br><br>a. <1%<br>b. 2 - 6%<br>c. 12 - 17%<br>d. 50 - 65% | The HEART score is a clinical tool used to estimate the risk of a major adverse cardiac event (MACE) within the next 6 weeks. It is calculated based on five factors, each scored from 0-2:<br><br>1. History (H): This patient's history is slightly suspicious. They have started to feel short of breath during exercise, which improved with rest. This could be indicative of exertional angina. Score: 1<br>2. ECG (E): T wave inversion or flattening is abnormal. Score: 2<br>3. Age (A): The patient is older than 65. Score: 2<br>4. Risk Factors (R): The patient is overweight and has a family history of heart disease (father had a heart attack in his early 60s). They are also on medication for blood pressure and cholesterol. Score: 2<br>5. Troponin (T): This data is not given. We'll assume it's 0 for the sake of this question.<br><br>Adding these together gives a HEART score of 7, which places the patient in the moderate |

| | | |
|---|---|---|
| | | risk category (score 4-6), corresponding to a 12 - 17% chance of MACE within 6 weeks. So, the answer is C. 12 - 17%.<br><br>However, please note that this calculation is not entirely accurate as we don't have the troponin levels. A blood test for troponin levels is critical for the definitive diagnosis, and the score could be higher or lower depending on that result. It is important to consider further diagnostic testing like stress testing or angiography based on the patient's clinical situation and risk tolerance. |

| Supplementary 5: Breakdown of Question Accuracy Results Before and After LLM Intervention | | | |
|---|---|---|---|
| **Question Number** | **Pre and post-intervention** | **Group A (White Male) N=25** | **Group B (Black Female) N=25** |
| **1** | How many answered Q1 correctly before intervention? | 3 (12%) | 3 (12%) |
| | How many answered Q1 correctly post-LLM intervention? | 11 (44%) | 14 (56%) |
| | Pre-Post Change | +8 (+32%) | +11 (+44%) |
| **2** | How many answered Q2 correctly before intervention? | 15 (60%) | 19 (76%) |
| | How many answered Q2 correctly post-LLM intervention? | 21 (84%) | 23 (92%) |
| | Pre-Post Change | +6 (+24%) | 4 (+16%) |
| **3** | How many answered Q3 correctly before intervention? | 17 (68%) | 19 (76%) |
| | How many answered Q3 correctly post-LLM intervention? | 20 (80%) | 19 (76%) |
| | Pre-Post Change | +3 (+12%) | 0 (0%) |
| **4** | How many answered Q4 correctly before intervention? | 12 (48%) | 21 (84%) |
| | How many answered Q4 correctly post-LLM intervention? | 14 (56%) | 24 (96%) |
| | Pre-Post Change | +2 (+8%) | +3 (+12%) |

| **Supplementary 6: Outcomes derived from statistical model** | | | |
|---|---|---|---|
| **Comparison** | **Difference in Score (Out of 4) (95% CI)** | **Standard error** | **P-value** |
| Group B (Black female Actor) vs Group A (White male actor) groups | +0.58 (0.20-0.96) | 0.19 | 0.00427 |
| Post intervention vs pre intervention | +0.74 (0.48-0.99) | 0.12 | $4.44 \times 10^{-7}$ |

| **Supplementary 7: Statistical Model Results of LLM intervention** | | | |
|---|---|---|---|
| **Covariate** | **Estimate** | **Std. Error** | **Test statistic** |
| Intercept | 1.88 | 0.1641 | < 2e-16* |
| Post-LLM intervention | 0.76 | 0.1817 | 0.000121* |
| Group-B (Black Female Vignette) | 0.60 | 0.2321 | 0.011470* |
| Post:Group-B Interaction | -0.04 | 0.2559 | 0.876923 |

| Supplementary 8: Post-task Survey Questions | |
|---|---|
| **Question No.** | **Questions** |
| 1-5 | We will use the NASA Task Load Index to measure the level of mental workload associated with completing the tasks today. In this section, you will report the mental workload when making a decision with the support of ChatGPT. What level of mental demand, temporal demand, performance, effort, and frustration did you feel today when using ChatGPT to complete tasks (1 = Lowest, 20 = Highest)?<br><br>**1. Mental Demand**:<br>How much thinking, deciding, or calculating was required to perform the task?<br><br>**2. Temporal Demand**:<br>How much time pressure did you feel while completing the task?<br><br>**3. Performance**:<br>How successful do you think you were in accomplishing the goals of the task?<br><br>**4. Effort**:<br>How hard did you have to work to accomplish your level of performance?"<br><br>**5. Frustration**:<br>From insecure and discouraged to secure and content, how did you feel during the task? |
| 6 | Have you used ChatGPT before today for any healthcare-related applications? (if yes, please elaborate for what purpose in the text box) |
| 7 | How likely is it that large language model (LLM) tools like ChatGPT will have a role to play in healthcare for clinical decision making? |
| 8 | What changes would you recommend making to large language model (LLM) tools like ChatGPT for use in healthcare for clinical decision making? |
| 9 | In what ways do you think that large language model (LLM) tools like ChatGPT could be most helpful to you? |
| 10 | How similar were the cases viewed today to cases you see in your day-to-day practice (in terms of complexity, demographics, information available, etc.)? |
| 11 | How similar was this task to the tasks you complete during a typical week as a clinician? (ie, diagnostic evaluation, deciding on management plan for patients etc) |

| Supplementary 9 - Likelihood of LLM Role in Clinical Decision Making | |
|---|---|
| **Likelihood of LLM Role in Clinical Decision Making** | **Count (n= 50)** |
| Very unlikely | 4 (8%) |
| Somewhat unlikely | 1 (2%) |
| Neither likely or unlikely | 0 (0%) |
| Somewhat likely | 12 (24%) |
| Very likely | 33 (66%) |
